# Do Not Attempt Resuscitation (DNAR) status in people with suspected COVID-19: Secondary analysis of the PRIEST observational cohort study

**DOI:** 10.1101/2021.01.23.21249978

**Authors:** Laura Sutton, Steve Goodacre, Ben Thomas, Sarah Connelly

## Abstract

**Background:** Cardiac arrest is common in people admitted with suspected COVID-19 and has a poor prognosis. Do Not Attempt Resuscitation (DNAR) orders can reduce the risk of futile resuscitation attempts but have raised ethical concerns.

**Objectives:** We aimed to describe the characteristics and outcomes of adults admitted to hospital with suspected COVID-19 according to their DNAR status and identify factors associated with an early DNAR decision.

**Methods:** We undertook a secondary analysis of 13977 adults admitted to hospital with suspected COVID-19 and included in the Pandemic Respiratory Infection Emergency System Triage (PRIEST) study. We recorded presenting characteristics and outcomes (death or organ support) up to 30 days. We categorised patients as early DNAR (occurring before or on the day of admission) or late/no DNAR (no DNAR or occurring after the day of admission). We undertook descriptive analysis comparing these groups and multivariable analysis to identify independent predictors of early DNAR.

**Results:** We excluded 1249 with missing DNAR data, and identified 3929/12748 (31%) with an early DNAR decision. They had higher mortality (40.7% v 13.1%) and lower use of any organ support (11.6% v 15.7%), but received a range of organ support interventions, with some being used at rates comparable to those with late or no DNAR (e.g. non-invasive ventilation 4.4% v 3.5%). On multivariable analysis, older age (p<0.001), active malignancy (p<0.001), chronic lung disease (p<0.001), limited performance status (p<0.001), and abnormal physiological variables were associated with increased recording of early DNAR. Asian ethnicity was associated with reduced recording of early DNAR (p=0.001).

**Conclusions:** Early DNAR decisions were associated with recognised predictors of adverse outcome, and were inversely associated with Asian ethnicity. Most people with an early DNAR decision survived to 30 days and many received potentially life-saving interventions.

**Registration:** ISRCTN registry, ISRCTN28342533, http://www.isrctn.com/ISRCTN28342533

## Introduction

In-hospital cardiac arrest is relatively common in patients with COVID-19 and often results in poor outcome. A multicentre cohort study from the United States [1] reported that 701/5019 (14.0%) critically ill patients with COVID-19 had in-hospital cardiac arrest, with 400/701 (57.1%) receiving CPR, and only 7% of these surviving to hospital discharge with normal or mildly impaired neurological status. Management of cardiac arrest in COVID-19 is further complicated by concerns about infection risk associated with aerosol-generating procedures and consequent risks to staff. [2]

These concerns have raised awareness about the need to consider do not attempt resuscitation (DNAR) decisions when patients are admitted to hospital with suspected COVID-19. An appropriately implemented DNAR decision can ensure that the patient’s wishes and best interests are addressed, while avoiding futile medical intervention.[3] However, concerns have been raised about inappropriate use of DNAR orders during the pandemic, [4] leading to the Care Quality Commission being asked to review their use in the United Kingdom (UK). [5]

Previous studies have estimated the prevalence of DNAR orders in patients admitted to hospital with community-acquired pneumonia [6-10] and sepsis,[11-14] and have attempted to identify factors associated with DNAR use, but we currently know very little about how DNAR orders have been used in people admitted with suspected COVID-19. The Pandemic Respiratory Infection Emergency System Triage (PRIEST) study was established to develop and evaluate triage tools for people presenting to hospital emergency departments with suspected COVID-19.[15] DNAR status was recorded to facilitate evaluation of triage tools in pre-specified subgroups. We present a post hoc secondary analysis of patients admitted with suspected COVID-19 that aims to describe their characteristics and outcomes according to their DNAR status and identify factors associated with recording of a DNAR decision.

## Methods

PRIEST was an observational cohort study of patients attending an emergency department (ED) in the UK with suspected COVID-19 infection during the first wave of the pandemic. We included patients if the assessing clinician recorded that the patient had suspected COVID-19 in the ED records or completed a standardised assessment form for suspected COVID-19 patients. The clinical diagnostic criteria for COVID-19 during the study were of fever (≥ 37.8°C) and at least one of the following respiratory symptoms, which must be of acute onset: persistent cough (with or without sputum), hoarseness, nasal discharge or congestion, shortness of breath, sore throat, wheezing, sneezing. We did not seek consent to collect data but information about the study was provided in the ED and patients could withdraw their data at their request. Patients with multiple presentations to hospital were only included once, using data from the first presentation identified by research staff.

We only included patients who were admitted to hospital after ED assessment because DNAR planning was considered unlikely to be routinely undertaken for discharged patients and would be limited to a minority of highly selected cases. We also only included adults (age ≥ 16 years) because previous analysis [15] showed that children with suspected COVID-19 had very low rates of confirmed COVID-19 or adverse outcome.

Baseline characteristics at presentation to the ED were recorded prospectively, using a standardised assessment form that doubled as a clinical record (Appendix 1: Standardised Data Collection Form), or retrospectively, through research staff extracting data onto the standardised form using the clinical records. Research staff collected follow-up data onto a standardised follow-up form (Appendix 2: Follow-up Form) using clinical records up to 30 days after presentation. This included recording whether a DNAR decision made at any time between initial presentation and follow-up, and if so, the date of the decision.

Patients who died or required respiratory, cardiovascular, or renal support were classified as having an adverse outcome. Patients who survived to 30 days without requiring respiratory, cardiovascular or renal support were classified as having no adverse outcome. Respiratory support was defined as any intervention to protect the patient’s airway or assist their ventilation, including non-invasive ventilation, or acute administration of continuous positive airway pressure. It did not include supplemental oxygen alone or nebulised bronchodilators. Cardiovascular support was defined as any intervention to maintain organ perfusion, such as inotropic drugs, or invasively monitor cardiovascular status, such as central venous pressure or pulmonary artery pressure monitoring, or arterial blood pressure monitoring. It did not include peripheral intravenous cannulation, or fluid administration. Renal support was defined as any intervention to assist renal function, such as haemofiltration, haemodialysis, or peritoneal dialysis. It did not include intravenous fluid administration.

We compared the characteristics and outcomes of those with a DNAR decision recorded on or before the day of ED assessment (early DNAR) to those with no DNAR recorded or a DNAR decision recorded at a later date (late/no DNAR). We categorised patients in this way on the assumption that patients with a late DNAR decision were likely to be systematically different from those with an early decision, with implementation of a late DNAR decision reflecting the response to intervention. Our categorisation was therefore based on a theoretical framework in which patient characteristics at admission could determine whether a DNAR decision was recorded at hospital admission, and recording of a DNAR decision at admission could then determine subsequent use of interventions.

We calculated a National Early Warning Score (2nd version, NEWS2) for adults, [16] to provide an overall assessment of acute illness severity on a score from zero to 20, based on respiratory rate, oxygen saturation, systolic blood pressure, heart rate, level of consciousness and temperature. We calculated a PRIEST COVID-19 clinical severity score, to provide an overall prediction of the risk of adverse outcome on a score from zero to 29, based on NEWS2, age, sex and performance status [17].

We also undertook multivariable logistic regression modelling to identify independent predictors of DNAR status. Variables were selected on the basis of clinical interest. Collinearity was observed between Glasgow Coma Scale (GCS) scores and consciousness, recorded as alert, responsive to verbal stimuli, responsive to pain or unconscious (AVPU). Missing AVPU data were imputed using GCS as follows, and GCS dropped from the list of predictors: GCS 15 = Alert, GCS 9-14 = Verbal, GCS 7-8 = Pain, and GCS 3-6 = Unresponsive. Continuous physiological predictors were categorised in accordance with NEWS2 risk categories [16] where the reference category denotes normal range and increasing category levels indicate increasing deviation from the norm. Data were analysed using SAS v9.4.

### Ethical approval

The North West - Haydock Research Ethics Committee (REC) gave a favourable opinion on the PAINTED study on 25 June 2012 (reference 12/NW/0303) and on the updated PRIEST study on 23rd March 2020. The Confidentiality Advisory Group of the Health Research Authority granted approval to collect data without patient consent in line with Section 251 of the National Health Service Act 2006. The REC approved a substantial amendment to undertake this secondary analysis on 7 January 2021.

## Results

The PRIEST study recruited 22484 patients from 70 EDs across 53 sites between 26 March 2020 and 28 May 2020, including 13997 adults admitted following ED assessment. Of these, 1249 had unknown DNAR status or timing, and were excluded from the analysis. The remaining 12748 were grouped into those with DNAR decisions made on or before the day of initial ED assessment (N=3929, 31%) and those with no DNAR or DNAR decisions made at a later date (N=8819).

Table 1 shows presenting characteristics for both groups. Patients with a DNAR decision recorded on or before their day of attendance tended to be older and have a higher prevalence of comorbidities, limited activity or self-care. They also tended to have a slightly higher respiratory rate, lower oxygen saturation, and lower Glasgow Coma Scale (GCS), and were less likely to be alert.

**Table 1.** Presenting characteristics for admitted adults with DNAR decisions in place by the end of ED assessment (N=3929) and adults with no DNAR or DNAR decision made later (N=8819)

| Characteristic | Statistic/level | Early DNAR | No/late DNAR |
| --- | --- | --- | --- |
| Age (years) | N | 3929 | 8819 |
|  | Mean (SD) | 79.4 (10.9) | 63.9 (17.6) |
|  | Median (IQR) | 81 (73,87) | 65 (52,78) |
| Sex | Missing | 42 | 78 |
|  | Male | 2027 (52.1%) | 4704 (53.8%) |
|  | Female | 1860 (47.9%) | 4037 (46.2%) |
| Ethnicity | Missing/prefer not to say | 601 | 1746 |
|  | UK/Irish/other white | 3102 (93.2%) | 6027 (85.2%) |
|  | Asian | 83 (2.5%) | 468 (6.6%) |
|  | Black/African/Caribbean | 75 (2.3%) | 289 (4.1%) |
|  | Mixed/multiple ethnic groups | 27 (0.8%) | 102 (1.4%) |
|  | Other | 41 (1.2%) | 187 (2.6%) |
| Presenting features | Cough | 2117 (53.9%) | 5514 (62.5%) |
|  | Shortness of breath | 2955 (75.2%) | 6678 (75.7%) |
|  | Fever | 1777 (45.2%) | 4688 (53.2%) |
| Comorbidities | No Chronic disease | 323 (8.2%) | 2012 (22.8%) |
|  | Heart Disease | 1620 (41.2%) | 2036 (23.1%) |
|  | Renal impairment | 739 (18.8%) | 843 (9.6%) |
|  | Steroid therapy | 160 (4.1%) | 258 (2.9%) |
|  | Asthma | 450 (11.5%) | 1446 (16.4%) |
|  | Diabetes | 1123 (28.6%) | 2034 (23.1%) |
|  | Active malignancy | 370 (9.4%) | 508 (5.8%) |
|  | Immunosuppression | 126 (3.2%) | 326 (3.7%) |
|  | Other chronic lung disease | 1219 (31%) | 1685 (19.1%) |
|  | Hypertension | 1721 (43.8%) | 3039 (34.5%) |
| Symptom duration (days) | N | 3396 | 7986 |
|  | Mean (SD) | 5.7 (6.8) | 7.5 (8.4) |
|  | Median (IQR) | 3 (1,7) | 5 (2,10) |
| Heart rate (beats/min) | N | 3867 | 8664 |
|  | Mean (SD) | 95.5 (23) | 97.4 (22) |
|  | Median (IQR) | 93 (80,109) | 96 (82,111) |
| Respiratory rate (breaths/min) | N | 3867 | 8625 |
|  | Mean (SD) | 25.5 (7.5) | 23.9 (6.9) |
|  | Median (IQR) | 24 (20,29) | 22 (19,28) |
| Systolic BP (mmHg) | N | 3834 | 8629 |
|  | Mean (SD) | 131.8 (27.8) | 134.1 (25.6) |
|  | Median (IQR) | 130 (112,149) | 132 (117,149) |
| Diastolic BP (mmHg) | N | 3813 | 8587 |
|  | Mean (SD) | 73.5 (17.1) | 77.4 (16.3) |
|  | Median (IQR) | 72 (62,84) | 77 (67,87) |
| Temperature (°C) | N | 3803 | 8576 |
|  | Mean (SD) | 37.1 (1.2) | 37.3 (1.2) |
|  | Median (IQR) | 37 (36.4,37.9) | 37.2 (36.5,38.1) |
| Oxygen saturation (%) | N | 3893 | 8741 |
|  | Mean (SD) | 92.6 (8) | 94.1 (6.8) |
|  | Median (IQR) | 95 (91,97) | 96 (93,98) |
| Glasgow Coma Scale | N | 2985 | 6643 |
|  | Mean (SD) | 13.9 (2.1) | 14.7 (1.3) |
|  | Median (IQR) | 15 (14,15) | 15 (15,15) |
| AVPU | Missing | 610 | 1009 |
|  | Alert | 2877 (86.7%) | 7486 (95.9%) |
|  | Verbal | 291 (8.8%) | 229 (2.9%) |
|  | Pain | 97 (2.9%) | 50 (0.6%) |
|  | Unresponsive | 54 (1.6%) | 45 (0.6%) |
| Performance status | Missing | 222 | 455 |
|  | Unrestricted normal activity | 453 (12.2%) | 4341 (51.9%) |
|  | Limited strenuous activity, can do light activity | 448 (12.1%) | 1160 (13.9%) |
|  | Limited activity, can self care | 923 (24.9%) | 1351 (16.2%) |
|  | Limited self care | 1153 (31.1%) | 993 (11.9%) |
|  | Bed/chair bound, no self care | 730 (19.7%) | 519 (6.2%) |
| NEWS2 score | N | 3911 | 8723 |
|  | Mean (SD) | 6.4 (3.3) | 5.1 (3.1) |
|  | Median (IQR) | 6 (4,9) | 5 (3,7) |
| PRIEST score | N | 3870 | 8645 |
|  | Mean (SD) | 12.5 (3.9) | 8.9 (4.1) |
|  | Median (IQR) | 12 (10,15) | 9 (6,12) |

Figures 1 compares the NEWS2 scores for the two groups and shows that those with early DNAR decisions tended to be more acutely unwell (median score 6 versus 5). Figure 2 compares the PRIEST COVID-19 clinical severity scores for the two groups and shows that those with early DNAR decisions were at a higher risk of adverse outcome (median score 12 versus 9, respectively indicating 38% versus 26% expected risk of a 30-day adverse outcome) [17].

**Figure 1.**
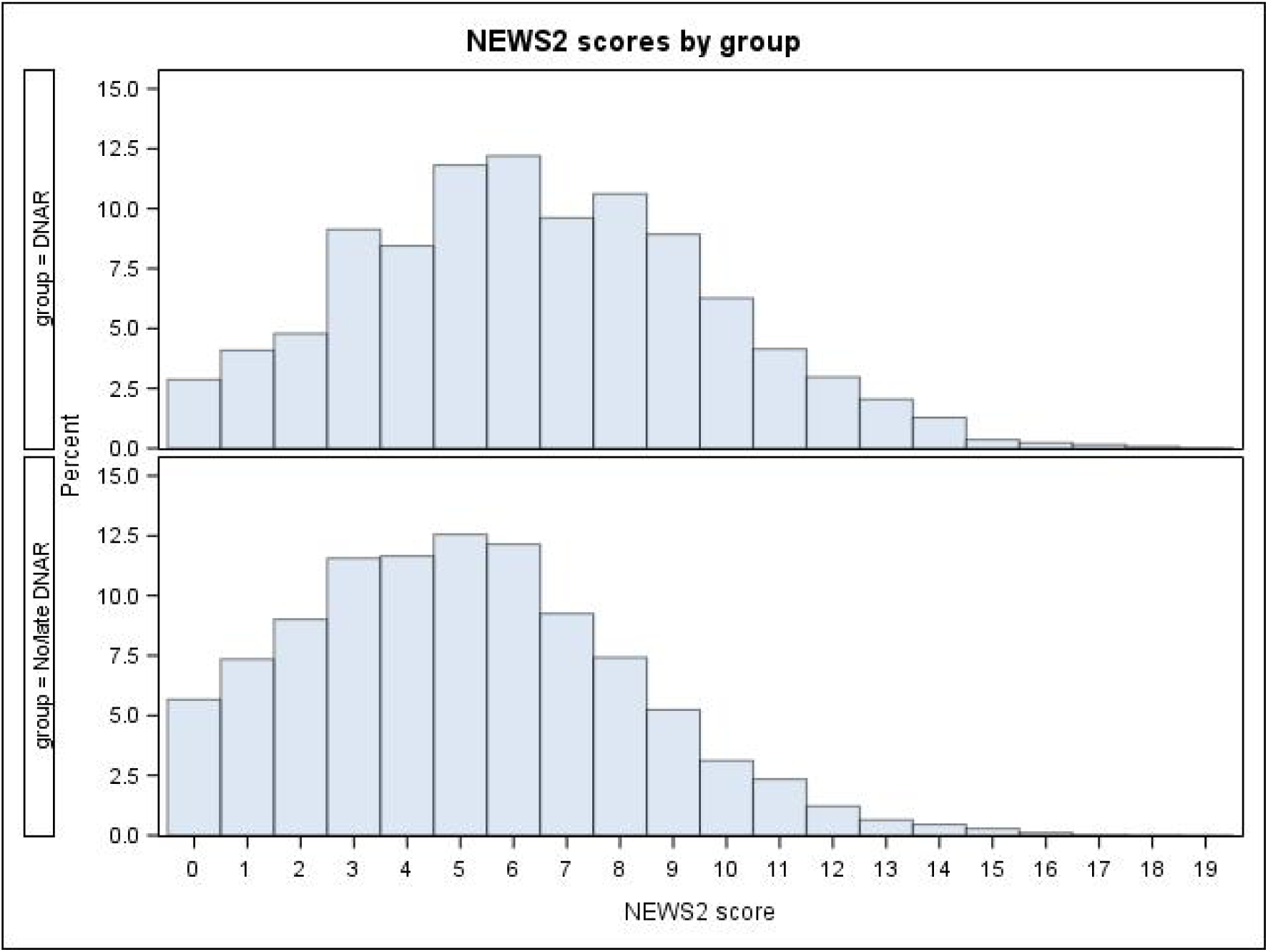
NEWS2 score distribution by DNAR status

**Figure 2.**
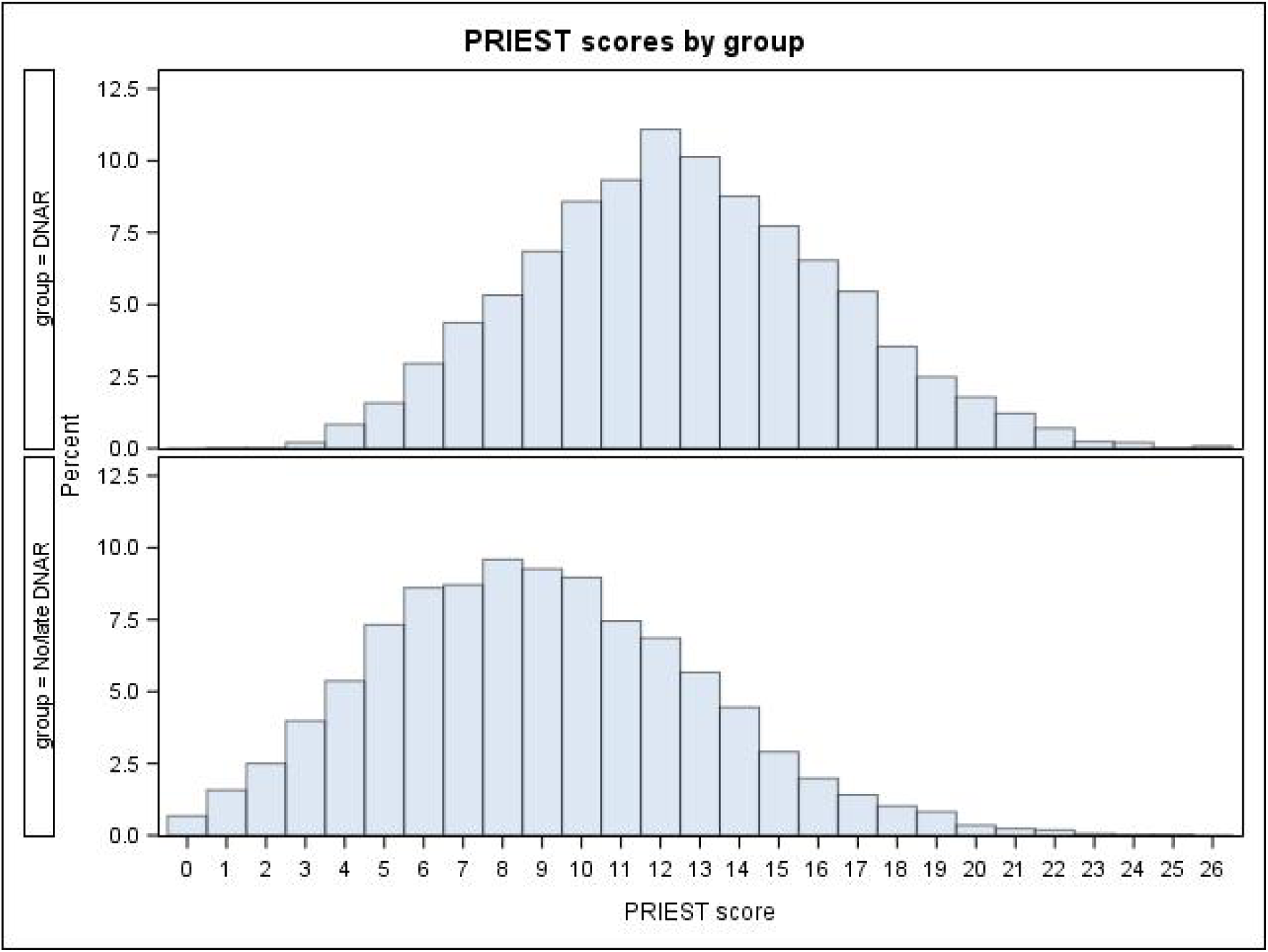
PRIEST score distribution by DNAR status

Table 2 gives location of admission, pathogen confirmation and adverse outcome data for the two groups. Patients with an early DNAR decision had a higher mortality rate but most (59.4%) survived to 30 days. They also had lower use of critical care and organ support, but a significant proportion (11.6%) received organ support. Table 2 shows the highest level of organ support received, according to a predefined hierarchy (corresponding to the order presented in the table). Patients with early DNAR decisions received a wide range of interventions, some at comparable rates to those with no or a late DNAR decision (e.g. non-invasive ventilation and high flow nasal oxygen).

**Table 2.**
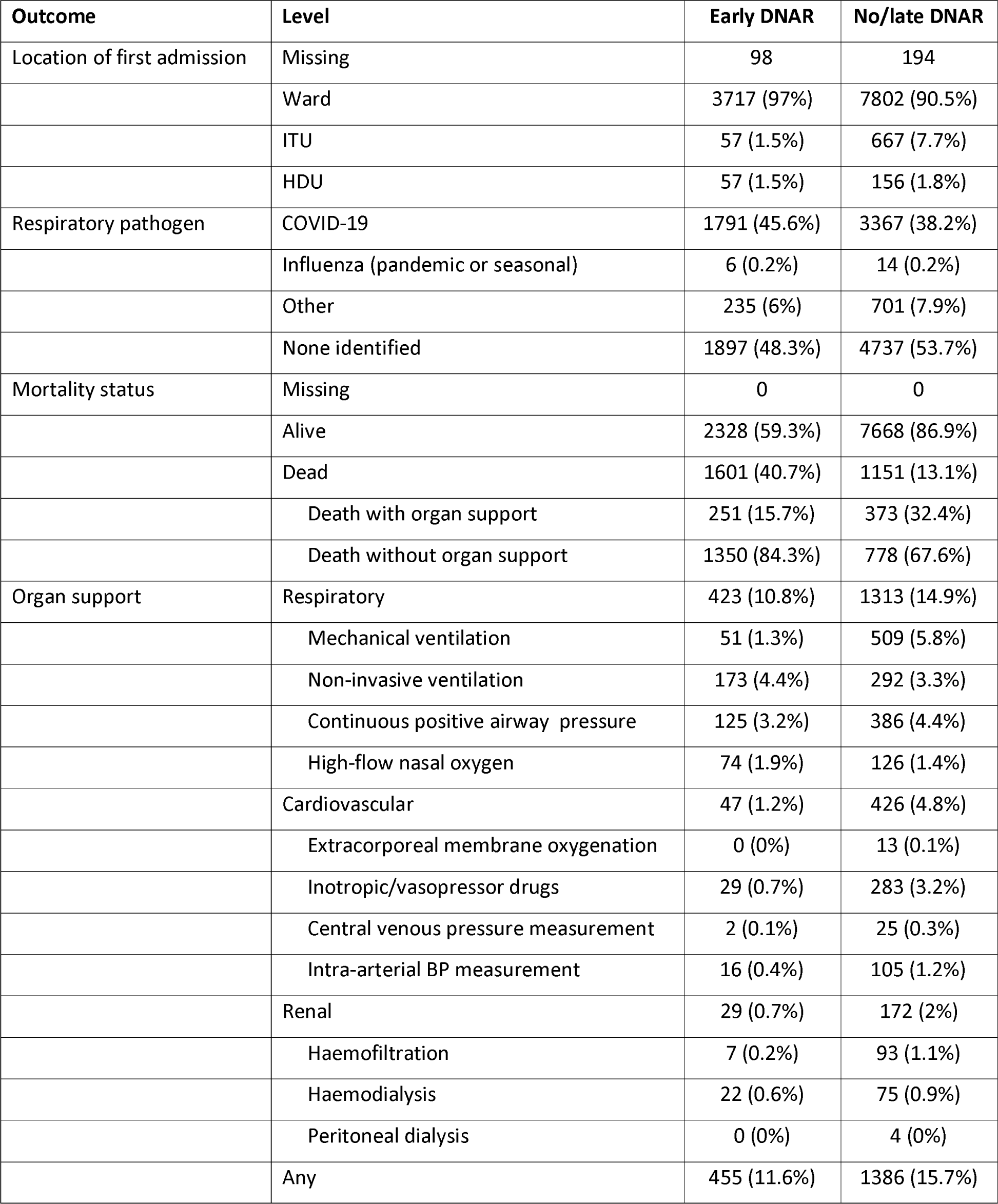
Outcome data for admitted adults with DNAR decisions in place by the end of ED assessment (N=3929) and adults with no DNAR or DNAR decision made later (N=8819)

Table 3 shows the results of the multivariable logistic regression model. Older age, active malignancy, chronic lung disease, limited performance status, abnormal heart rate, abnormal respiratory rate, lower oxygen saturation, and lower alertness were all associated with increased use of early DNAR. Asian ethnicity was associated with a lower use of early DNAR.

**Table 3.**
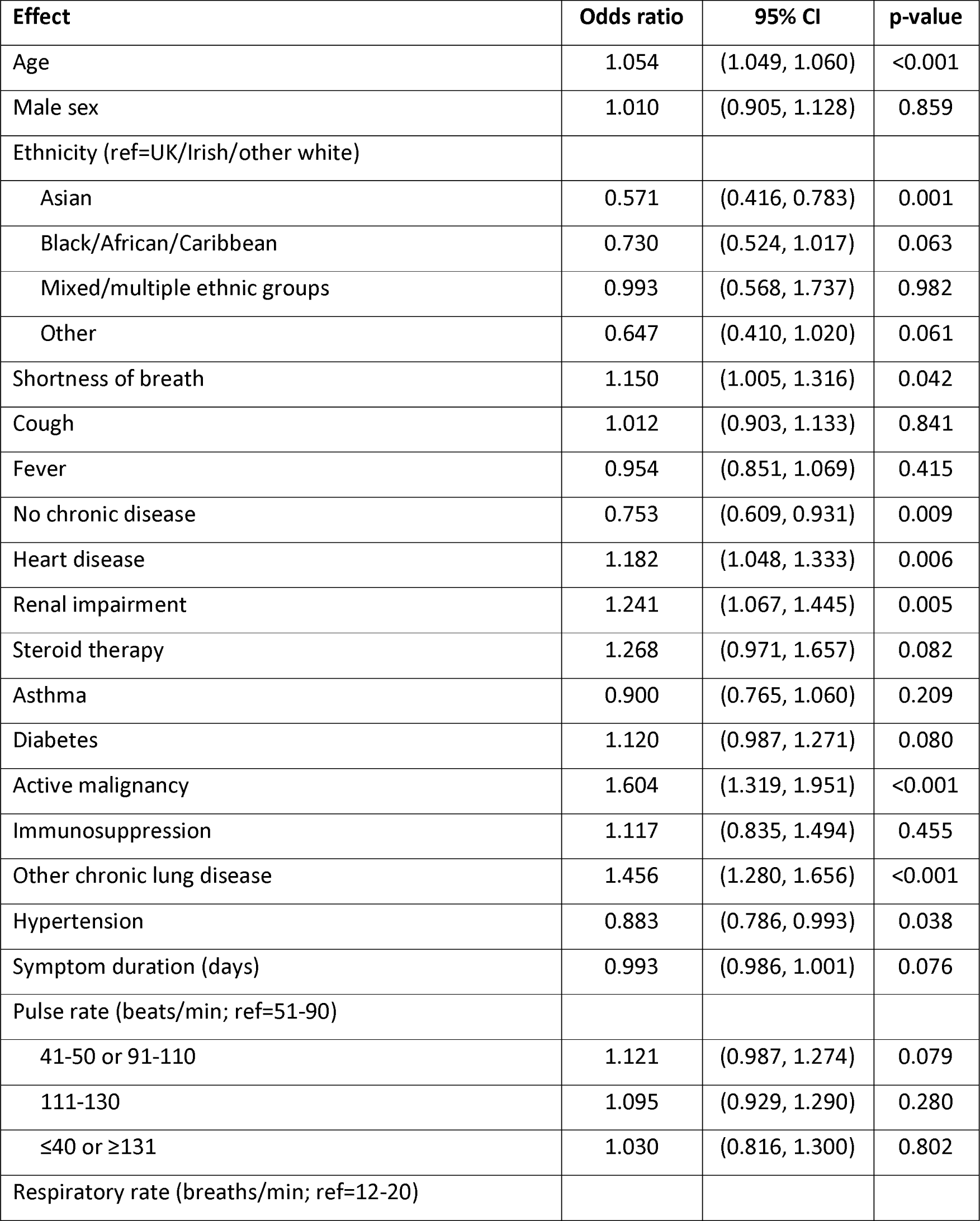
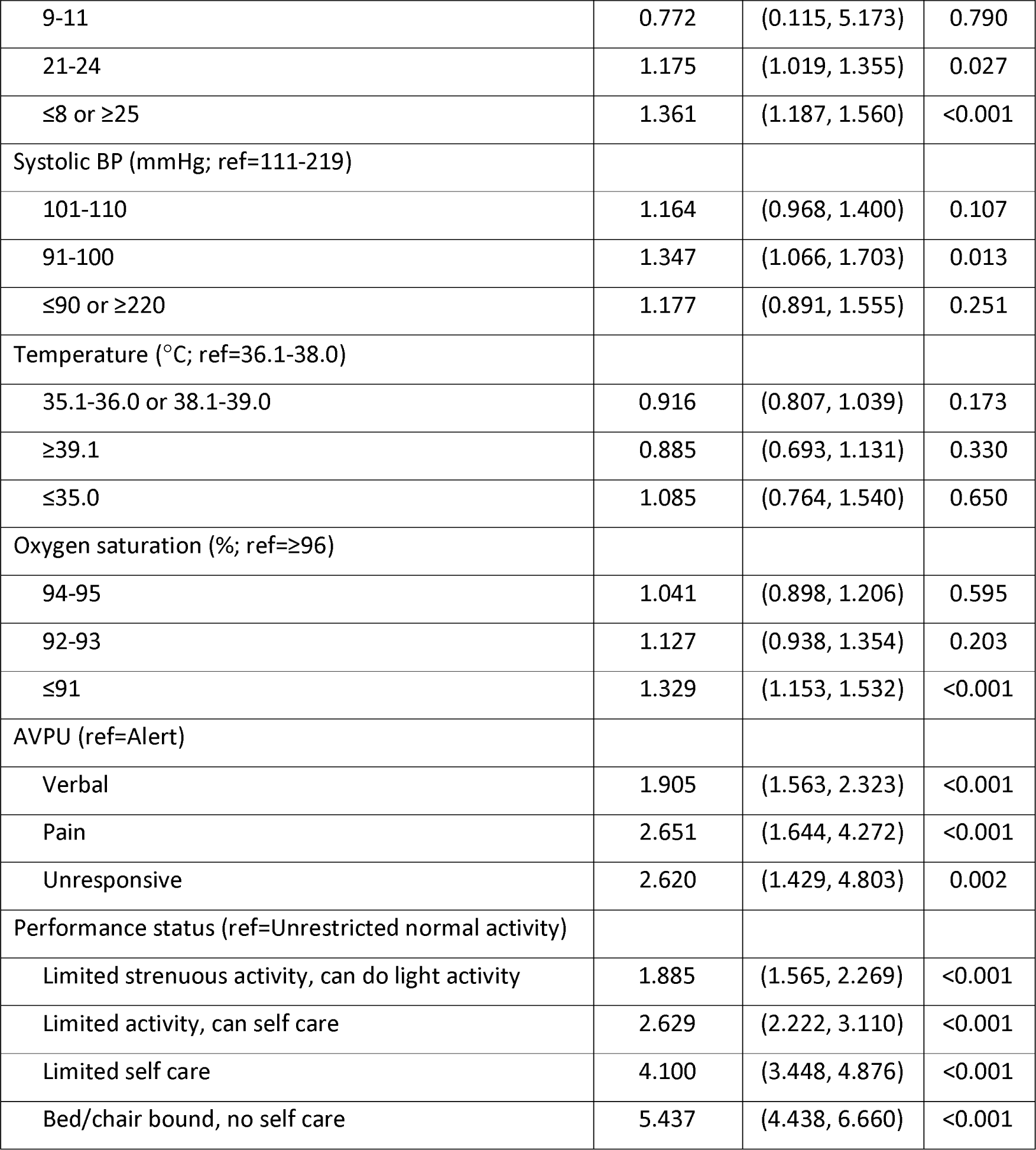
Multivariable analysis of predictors of early DNAR use

## Discussion

We found that 31% of adults admitted to hospital with suspected COVID-19 during the first phase of the pandemic had a DNAR decision recorded on or before their day of attendance, after excluding those who could not be classified. Most patients (59.4%) with an early DNAR decision survived to 30 days and 11.6% received some form of organ support. These findings show that potentially life-saving treatments were provided to a significant proportion of people, potentially addressing concerns that DNAR decisions may be conflated with ‘do not provide active treatment’.[18] The use of invasive intervention, particularly mechanical ventilation, in people with a DNAR decision was an unexpected finding. Contact with participating site investigators suggested that this could be explained by use of the ReSPECT process in discussions about resuscitation, in which the patient is encouraged to explicitly indicate which treatments they want in a future situation where they are unable to make or express choices.[19] The ReSPECT process therefore allows patients to consent to mechanical ventilation but decline cardiopulmonary resuscitation if it is subsequently required.

Older age, active malignancy, chronic lung disease (excluding asthma), and lower performance status were associated with increased use of early DNAR, whereas Asian ethnicity was associated with decreased use. Patients with early DNAR status tended to be more acutely ill, with higher NEWS2 scores. Abnormal respiratory rate, lower oxygen saturation, and lower alertness were associated with increased use of early DNAR.

Our findings suggest a higher rate of DNAR use (31%) than identified in previous studies of similar conditions. Studies of patients admitted with community-acquired pneumonia reported rates of DNAR use ranging from 13% to 29%, [6-10] while studies in severe sepsis or septic shock reported rates ranging from 9% to 20%. [11-14] DNAR decisions in these studies were associated with older age, but conflicting findings were reported around the use of invasive procedures. Sakari *et al* [11] and Bradford *et al* [12] reported that DNAR orders were associated with lower rates of invasive procedures, while Powell *et al* [13] reported no difference, and Huang *et al* [14] reported a higher rate of arterial or central venous cannulation in those with a DNAR order.

We found an association between Asian ethnicity and decreased use of early DNAR status compared to White ethnicity. The odds ratio for Black/African/Caribbean ethnicity also suggested decreased use but was not significant. Previous studies from the United States have shown less use of DNAR decisions among African-American, Asian, and Hispanic patients, [20,21,22] and Black patients tend to receive more life-prolonging treatment at end of life care.[23] A systematic review of end of life decisions for people from ethnic minority groups suggested that Hispanic and African American people had advance care plans documented less often, citing religious coping and spirituality as factors.[24] A scoping review of culturally- and spiritually-sensitive end-of-life care highlighted a multitude of factors influencing end-of-life care and subsequent experiences by culturally- and spiritually-diverse groups.[25] Further research of DNAR decisions in relation to ethnicity is clearly required.

Studies of DNAR use in COVID-19 are currently limited. Alhatem *et al* [26] analysed 1270 patients admitted across two hospitals with COVID-19, of whom 750/1270 (59%) had a DNAR order at admission, and 570/750 (76%) of these died. Age over 60, male sex, and comorbidities were associated with DNAR at admission. Coleman *et al* [27] examined records of DNAR decisions at a single centre from 2017 to 2020 and showed an increased rate of DNAR use during pandemic, with patients tending to be younger and have fewer comorbidities. It is unclear whether these findings reflect an increased overall need for DNAR decisions during the pandemic or increased willingness to use DNAR decisions in COVID-19. Our findings suggest that a relatively high rate of DNAR use in suspected COVID-19 may contribute to an increased rate of DNAR use during the pandemic.

This study was based on a large representative sample of adults admitted with suspected COVID-19, but has a number of limitations. DNAR decisions were recorded to facilitate subgroup analyses addressing the primary purpose of the study rather than addressing the aims of this secondary analysis. We were unable to include 1249/13997 (9%) cases because data were missing or uncertain regarding the use or timing for DNAR. Our categorisation on the basis of timing of DNAR decision and assumption that later DNAR decisions are qualitatively different to early decisions could be challenged. We did not collect any detailed data to allow us to explore the reasons behind DNAR decisions, so we are unable to offer explanations for the associations identified in our analysis. Our suggestion that the ReSPECT process could explain the use of invasive interventions in people with a DNAR decision is based on informal contacts and requires further research. The use of the ReSPECT process could also undermine our rationale for categorising DNAR decisions as early versus late or no decision, and suggests a complex relationship between DNAR decisions and subsequent interventions.

In conclusion, we found that many patients with an early DNAR decision went on to receive life-saving interventions and most survived to 30 days. Early DNAR decisions were associated with older age, lower performance status, active malignancy, chronic lung disease and severe illness, as indicated by physiological parameters. We found some evidence of an association between ethnicity and DNAR status that requires further research.

## Supporting information

Appendix 1: Data Collection Form

Appendix 2: Follow-up Form

Appendix 3: Study research team

Appendix 4: Study Steering Committee

Appendix 5: Site research teams

Appendix 6: CTRU acknowledgements

## Data Availability

Anonymised data are available from the corresponding author upon reasonable request

## Competing interests

All authors have completed the ICMJE uniform disclosure form at www.icmje.org/coi_disclosure.pdf and declare: grant funding to their employing institutions from the National Institute for Health Research; no financial relationships with any organisations that might have an interest in the submitted work in the previous three years; no other relationships or activities that could appear to have influenced the submitted work.

## Contributor and guarantor information

SG conceived and designed the study. BT oversaw data acquisition. LS analysed the data. SC undertook literature searches. SG, LS, BT and SC interpreted the data. All authors contributed to drafting the manuscript. SG is the guarantor of the paper. The corresponding author attests that all listed authors meet authorship criteria and that no others meeting the criteria have been omitted.

## Acknowledgements

We thank Katie Ridsdale for clerical assistance with the study, Erica Wallis (Sponsor representative), all members of the PRIEST Research Team, (Appendix 3), all members of the Study Steering Committee (Appendix 4) and the site research teams who delivered the data for the study (Appendix 5), and the research team at the University of Sheffield past and present (Appendix 6).

## Data sharing

Anonymised data are available from the corresponding author upon reasonable request (contact details on first page).

## Role of the funding source

The PRIEST study was funded by the United Kingdom National Institute for Health Research Health Technology Assessment (HTA) programme (project reference 11/46/07). The funder played no role in the study design; in the collection, analysis, and interpretation of data; in the writing of the report; and in the decision to submit the article for publication. The views expressed are those of the authors and not necessarily those of the NHS, the NIHR or the Department of Health and Social Care.

## Notes

### Competing Interest Statement

All authors declare grant funding to their employing institution from the National Institute for Health Research.

### Clinical Trial

http://www.isrctn.com/ISRCTN28342533

### Clinical Protocols

http://www.isrctn.com/ISRCTN28342533

https://www.sheffield.ac.uk/scharr/research/centres/cure/priest

https://www.fundingawards.nihr.ac.uk/award/11/46/07

### Summary of Updates

The abstract conclusion has been amended to specify that the association with Asian ethnicity was an inverse association. We have also uploaded the appendices.

