## Appendix 3: Study research team for "Do Not Attempt Resuscitation (DNAR) status in people with suspected COVID-19: Secondary analysis of the PRIEST observational cohort study"

**Appendix 3: The PRIEST study research team**

| **Name** | **Study Role** | **Role** | **Affiliation** |
| --- | --- | --- | --- |
| Ben Thomas | Study manager (lead) | Study Manager | University of Sheffield |
| Katie Biggs | CTRU over sight | Assistant Director | University of Sheffield |
| Steve Goodacre | Chief Investigator | Professor of Emergency Medicine | University of Sheffield |
| Carl Marincowitz | Clinical co-investigator | Clinical Lecturer in Emergency Medicine | University of Sheffield |
| Ellen Lee | Senior statistician | Statistician | University of Sheffield |
| Laura Sutton | Statistician | Statistician/Research Associate | University of Sheffield |
| Matthew Burnsall | Statistician | Statistician | University of Sheffield |
| Mike Bradburn | Senior Statistician | Senior Medical Statistician | University of Sheffield |
| Simon Waterhouse | Data Management | Lead Data Specialist | University of Sheffield |
| Richard Simmonds | Data Management | Data Management/Information Systems Officer | University of Sheffield |
| Jose Schutter | Research Assistant | Research Assistant | University of Sheffield |
| Sarah Connelly | Research Assistant | Research Assistant | University of Sheffield |
| Elena Sheldon | Research Assistant | Research Assistant | University of Sheffield |
| Jamie Hall | Research Assistant | Research Assistant | University of Sheffield |
| Emma Young | Research Assistant | Research Assistant | University of Sheffield |
| Ian Maconochie | Project Management Group, paediatric emergency medicine | Consultant in Paediatric Emergency Medicine / Associate Medical Director | Imperial College Healthcare NHS Trust |
| Andrew Lee | Project Management Group, public health | Reader of Global Public Health | University of Sheffield |
| Darren Walter | Project Management Group, emergency medicine | Clinical Senior Lecturer / Consultant in Emergency Medicine | Manchester University NHS Foundation Trust |
| Andrew Bentley | Project Management Group, critical care and respiratory medicine | Consultant in ICM & Respiratory Medicine | Manchester University NHS Foundation Trust, Wythenshawe Hospital |
| Chris Fitzimmons | Project Management Group, paediatric emergency medicine | Consultant in Paediatric Emergency Medicine | Sheffield Children's NHS Foundation Trust |
| Fiona Lecky | Project Management Group, emergency medicine | Clinical Professor in Emergency Medicine | University of Sheffield |
| Tim Harris | Project Management Group, emergency medicine | Professor of Emergency Medicine | Barts Health NHS Trust |
| Kirsty Challen | Project Management Group, emergency medicine | Consultant in Emergency Medicine | Lancashire Teaching Hospitals NHS Foundation Trust |
